# Unsupervised Extractive Summarization of Psychedelic User Experience Reports

**DOI:** 10.1101/2025.08.22.25334176

**Authors:** Shahidul Islam, Sakib Salam, Md Nahid Hasan

## Abstract

Contemporary psychedelic research highlights the value of user experience reports, yet their verbose, subjective nature poses challenges for clinical utility. This is the first study to pioneer unsupervised automatic text summarization of psychedelic user experience reports, a domain where no human-annotated reference summaries exist. To address this gap, we developed a custom scoring function that integrates semantic coverage, narrative coherence, and a novel experiential preservation metric, enabling effective model training and hyperparameter tuning. We utilized three established extractive methods: LexRank, LSA with HDBSCAN clustering, and SBERT with Maximal Marginal Relevance, on 1,200 reports involving LSD, psilocybin, and DMT. Using GPT-4 as a calibrated rater under a structured rubric, supplemented by TOPSIS aggregation, results showed LexRank achieving the highest overall balance with SBERT excelling in content coverage and experiential depth but lagging in coherence. Our findings revealed trade-offs between content richness and narrative fluency, with performance varying across substance types due to differences in narrative structure and phenomenology. Limitations included reliance on extractive methods, lack of reference data, and sensitivity to scoring design. Future work should extend to abstractive methods, alternative weighting schemes, and expert adjudication to develop clinically usable summarization systems for psychedelic science.

## 1 Introduction

### 1.1 Background

The renaissance of clinical research into psychedelics marks a significant paradigm shift in psychiatry and mental health care, characterized by a turbulent and dynamic timeline of scientific exploration and societal attitudes. Psychedelic compounds such as LSD, psilocybin, and mescaline were first investigated intensively in the 1950s and 1960s, when early clinical studies suggested promise for treating alcoholism, anxiety, and existential distress in terminal illness [1, 2]. However, widespread abuse, growing political concern, and the U.S. Controlled Substances Act of 1970 led to an abrupt research moratorium [3].

After decades of dormancy, psychedelic science reemerged in the late 1990s and early 2000s, catalyzed by work at Johns Hopkins, Imperial College London, and other prominent research hubs. Modern trials employ rigorous methodologies, revealing that psilocybin can produce substantial and enduring reductions in depressive symptoms, even among treatment-resistant patients [4]. Likewise, MDMA and 5-MeO-DMT assisted therapy has yielded remarkable improvements in PTSD symptoms [5, 6]. These recent findings have made psychedelics a key focus of modern psychiatric advancements breaking the barrier of pre-existing social norms..

Parallel to clinical trials, an expansive body of first-person trip narratives exists online via repositories like Erowid, Reddit, and PsychonautWiki. In contrast to formal clinical records, these user experience reports are often verbose narration filled with highly unstructured granular details, vivid emotional description, and subjective insights that reflect the complex, personal nature of psychedelic experiences. These seemingly story-like reports are actually invaluable for generating hypotheses, identifying risks, and understanding long-term effects of psychedelics. However, turning these detailed, unstructured narratives into concise, clinically relevant summaries utilizing Natural Language Processing techniques is quite challenging, which requires a balance of clarity, coherence, preservation as well as medical usefulness.

### 1.2 Prior Works and Research Gap

Recent computational work has applied NLP to these narratives, but primarily for classification, linguistic profiling, and predictive modeling, sentiment analysis rather than structured summarization. For instance, Noah et al. conducted a large-scale study of 103 psychoactive substances using embedding-based models to identify variability in visual effects, such as movement, color, and patterns [7]. Their work demonstrated cross-substance distinctions but remained narrowly focused on perceptual categories. Al-Imam et al. analyzed over 2,100 reports of LSD and psilocybin, applying BERT, RoBERTa, and VADER methods to identify emotional polarity and introspective themes [8]. Sentiment analysis of this study showed that VADER produced more polarized results, while RoBERTa offered cautious and more accurate classifications. Lexicon analysis revealed that psilocybin (mushroom) reports often focused on introspection and altered time perception, while LSD reports emphasized cognitive disturbances. On the other hand, Biba and O’Shea analyzed a large corpus of reddit reports pertaining to the discussion of psychedelics using NLP infrastructure to find out public sentiment polarization towards the consumption [9].

Other studies highlight the potential of computational text analysis for understanding psychedelic experiences. Tagliazucchi emphasized language as a “window into altered consciousness,” demonstrating semantic and structural markers of psychedelic states with predictive value for therapeutic outcomes [10]. Hase et al. extended this line of work by profiling distinct linguistic signatures across psychedelics and antidepressants, finding substance-specific language markers (e.g., MDMA as emotional, DMT as analytical) [11]. Furthermore, Cox et al. used topic modeling of 1,141 reports to predict substance reduction outcomes, showing clinical predictive utility but focusing on behavioral prediction rather than experiential condensation [12].

Nonetheless, these studies leave a significant gap in systematic summarization of full-length, phenomenology-rich trip reports. Numerous review works [13–15] divide the Automatic Text Summarization (ATS) techniques into two broad category: extractive and abstractive methods. Extractive summarization preserves original wording by selecting salient sentences, while abstractive approaches generate paraphrased digests. Clinical NLP has produced robust frameworks for summarizing medical records and patient notes, focusing on symptoms, diagnoses, and treatments. However, psychedelic trip reports differ fundamentally. They are subjective, nonlinear, and often metaphorical. Moreover, conventional evaluation frameworks often rely on reference summaries or objective clinical criteria, which are absent in this niche criterion.

Therefore, the non-existence of effective ATS and reference summaries for psychedelic trip reports underscores the urgent need for innovative summarization methods tailored to these unique experiences.

### 1.3 Research Aim

With this study, we aimed to address the challenges of summarizing psychedelic trip reports by developing a clinically oriented summarization pipeline that reduces narrative length by approximately 65-75% while preserving essential experiential and clinical content. We developed a custom scoring function for model training and hyperparameter tuning pertaining to this unique research in absence of gold-standard reference data. Additionally, we compared those extractive summarization models across narratives of LSD, psilocybin, and DMT, evaluating how their distinct phenomenological structures influence model performance. We believe that generated summaries from our research can reduce clinicians’ reading burden while retaining key experiential and risk-related information to support therapeutic applications.

## 2 Methodology

### 2.1 Data Collection and Preprocessing

We collected 400 narrative reports each for DMT, psilocybin, and LSD (total 1,200 reports) from the Erowid experience archive, with permission from the copyright holders. Previous research has demonstrated the value of Erowid as a corpus for computational and clinical investigations. For instance, Sanz et al. (2018) analyzed Erowid reports to identify symptom dimensions of hallucinogen use, while Swogger et al. (2015) conducted a qualitative thematic analysis of kratom user experiences hosted on Erowid.org, highlighting both therapeutic and adverse outcomes [16, 17]. Following these precedents, we constructed a balanced psychedelic-specific dataset stratified by substance. We excluded reports shorter than 500 words or longer than 2,500 words to ensure narrative depth and analytical consistency.

All reports were already anonymous and pseudonymized by Erowid moderators and users. Nonetheless, we conducted an additional named-entity recognition (NER) pass to remove any residual personal identifiers, ensuring full de-identification. To prepare the narratives for analysis, a systematic text cleaning and preprocessing pipeline was implemented. Structural metadata (e.g., “BODY WEIGHT,” “Dosage,” “Exp Year”) was truncated using regular expressions, retaining only the experiential content. Redundancy was reduced by eliminating duplicate sentences within each report. Text normalization proceeded in several stages: contractions were expanded (e.g., “don’t” to “do not”), clipped leading contractions (“’ve,” “’d”) were mapped to full grammatical forms (“i have,” “i would”), and parenthetical or bracketed insertions were removed. Punctuation was standardized by replacing em/en dashes and ellipses with commas, and collapsing repeated punctuation marks. Encoding artifacts (mojibake) were corrected via rule-based replacement of malformed Unicode sequences (e.g., “â€™” to “’”). We further applied case normalization, split camel-Case expressions, and enforced consistent whitespace. In addition to text cleaning, demographic metadata were extracted where available. Reported ages were parsed from the “Age at time of experience” field, and weights expressed in pounds or stone were converted to kilograms for consistency.

Finally, we conducted descriptive analyses of demographic and textual distributions across substances. We visualized gender proportions with pie charts and depicted age and weight distributions with histograms and strip plots. Moreover, we assessed narrative length variations via word-count histograms. These preprocessing steps ensured the dataset was structurally consistent, demographically harmonized, and analytically robust for subsequent model training, parameter tuning, and evaluation. In our workflow, we divided the corpus into ~75% for model training, hyperparameter tuning and ~25% for evaluation, stratified uniformly across LSD, psilocybin, and DMT to preserve substance-specific balance. We trained and tuned the models on the ~75% subset using an Optuna-based parameter search (Akiba et al., 2019), and then applied the optimal configurations to the held-out ~25% subset to generate summaries. These summaries were subsequently evaluated using the LLM rubric framework described afterwards. This framework leverages large language models as evaluators, a scalable alternative to human judgment in NLP task [18, 19].

### 2.2 Models and Hyperparameter Tuning

We evaluated three unsupervised extractive summarization approaches. The first was LexRank, a graph-based centrality algorithm. LexRank represents sentences as nodes in a similarity graph, with edges weighted by TF–IDF cosine similarity [20]. Low-similarity edges are pruned, and sentence salience is computed through eigenvector centrality. The top-ranked sentences are finally selected according to a target compression ratio.

The second approach combined Latent Semantic Analysis (LSA) with HDBSCAN clustering to identify semantically coherent sentence groups. LSA reduces TF–IDF sentence vectors into a lower-dimensional latent space using singular value decomposition (SVD) [21]. Sentences are subsequently grouped using HDBSCAN, a hierarchical density-based clustering algorithm [22]. This hybrid approach balances topical representativeness with phenomenological richness. The third approach employed Sentence-BERT (SBERT) with Maximal Marginal Relevance (MMR) for embedding-based sentence selection. SBERT generates dense semantic embeddings using transformer encoders [23]. MMR iteratively selects sentences by balancing document relevance and diversity [24]. Relevance is computed against a document embedding, while diversity penalizes redundancy among selected sentences. Additional tuning adjusts position bias and similarity thresholds. This framework enforces both semantic fidelity and experiential diversity, making it particularly well-suited for the multi-faceted nature of psychedelic narratives.

All three models were tuned using Optuna with a Tree-structured Parzen Estimator (TPE) sampler and Median Pruner, allowing efficient exploration of hyperparameter spaces and early stopping of unpromising trials. The optimization objective was a custom composite score balancing semantic fidelity, phenomenological preservation, and narrative coherence, as expressed by:

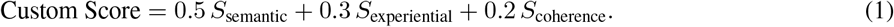

We computed semantic similarity as cosine similarity between the summary and source (TF–IDF for LexRank and LSA+HDBSCAN; SBERT embeddings for SBERT+MMR). To evaluate experiential preservation, we curated a lexicon of terms spanning six distinct domains, obtaining initial vocabulary with prior psychometric work on altered states of consciousness by Studerus et al.[25]. Then we expanded it using GPT-4 to capture synonyms and phrase variants. We measured preservation as the proportion of experiential terms in the source that appeared in the summary. To assess sentence coherence, we averaged pairwise cosine similarity between consecutive summary sentences. A representative excerpt of our experiential lexicon with respective domains is shown in Table 1, with the complete list provided in Appendix A. Furthermore, the hyperparameter search space of every model is summarized in Table 2.

**Table 1:** Representative Experiential Terms by Category. The full lexicon is provided in Appendix A.

| Category | Sample Terms |
| --- | --- |
| Emotional | joy, fear, euphoria, sadness, compassion |
| Sensory | visual, sound, fractal, shimmering, synesthesia |
| Cognitive | thought, clarity, ego, looping, interpretation |
| Physical | body, tingling, nausea, heartbeat, chills |
| Mystical | unity, transcend, ego death, nirvana, divine |
| Temporal | onset, peak, comedown, wave, time distortion |

**Table 2:**
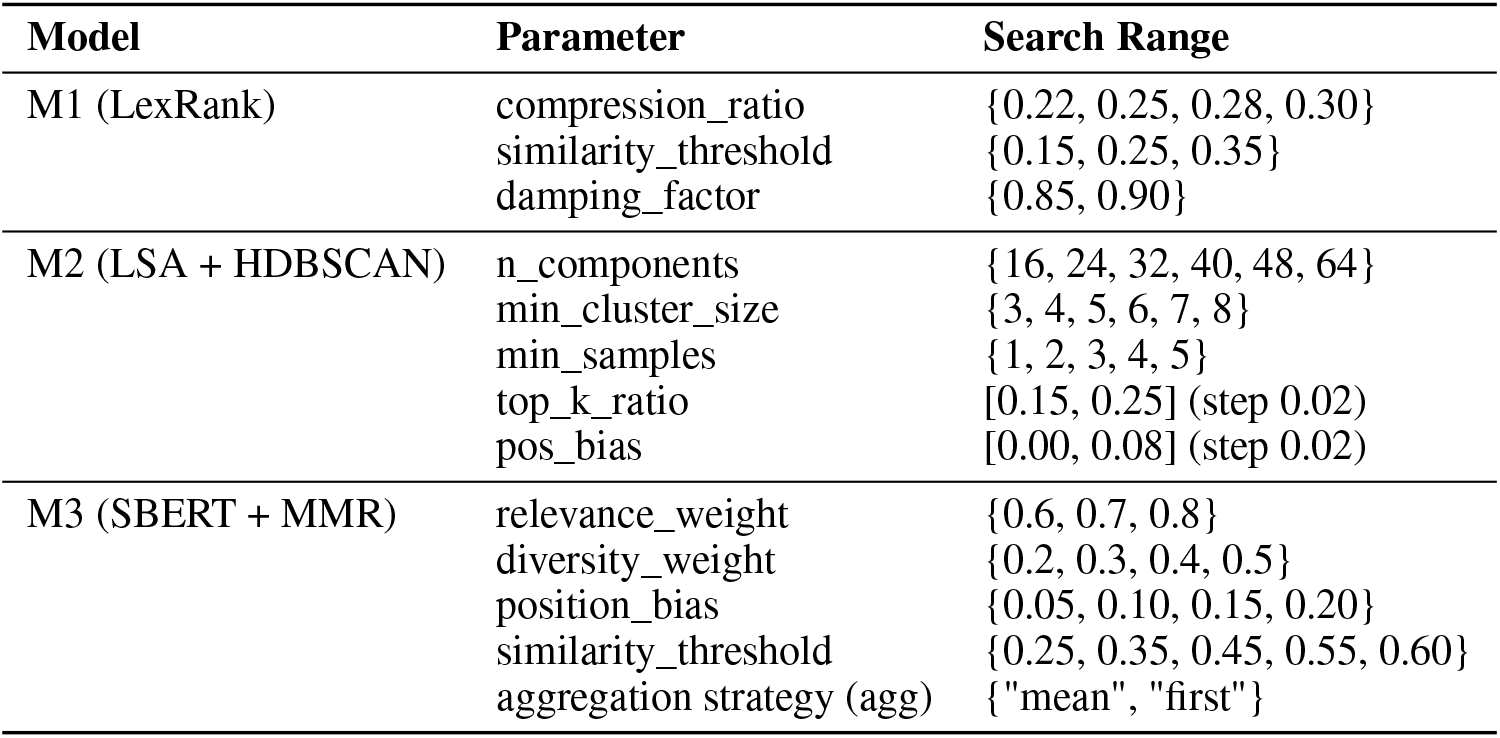
Hyperparameter Configuration and Tuning Ranges for Each Model.

### 2.3 LLM Evaluation

We evaluated the extracted summaries with optimal hyper-parameters using GPT-4 acting as a calibrated expert rater under a structured rubric. The exact prompt given in Table 3 is reproduced to ensure transparency and reproducibility.

**Table 3:** LLM Evaluation Prompt (GPT-4) used for psychedelic trip report summarization.

| LLM Evaluation Prompt Template for Psychedelic Trip Report Summarization |  |
| --- | --- |
| **Instruction:** | You are a clinical researcher specializing in psychedelic-assisted therapy. Evaluate the summary against the original trip report using strict, evidence-based criteria. |
| **Original Report:** | [report_text] |
| **Generated Summary:** | [summary] |
| **Evaluation Criteria:** | 1. Content Coverage (1–5): Does the summary capture the key experiences, effects, and important details?<br>2. Narrative Coherence (1–5): Does the summary flow logically and maintain fluency?<br>3. Clinical Relevance (1–5): Would this summary be useful for healthcare providers or researchers?<br>4. Compression Quality (1–5): Is the length appropriate while preserving essential information?<br>5. Experience Preservation (1–5): Are the subjective, experiential aspects adequately represented? |
| **Overall Score (1–5):** |  |
| **Brief Justification:** | [2–3 sentences explaining the rating] |

## 3 Results

Figure 1 illustrates the distribution of word counts and demographic attributes across psilocybin, LSD, and DMT reports. Word counts varied substantially across substances. LSD and psilocybin reports showed relatively normal distributions with peaks around 1,200–1,500 words, whereas DMT reports tended to be shorter, concentrated mainly between 500–1,000 words. Gender distribution was consistently male-dominated (79.5% for psilocybin, 76.2% for LSD, and 83.2% for DMT), with female contributions comprising less than one-fifth of the dataset. The age profiles revealed that the majority of participants fell within the 15–30 age range, with a small fraction of users showing slightly more spread into older cohorts. Weight distributions were broadly consistent across substances, although LSD users showed least outliers within the weight range.

**Figure 1.**
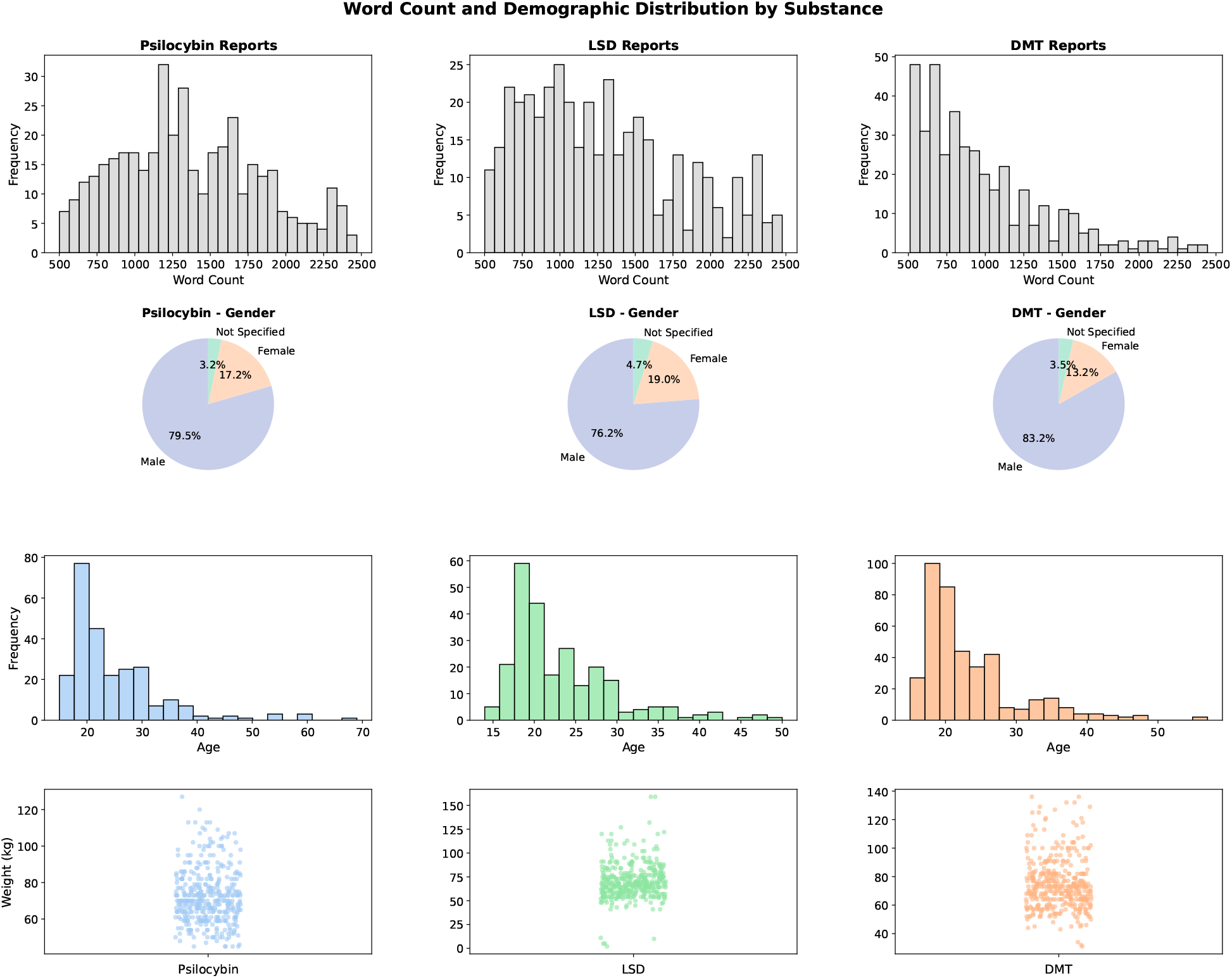
Distribution of word counts and demographics across substances (psilocybin, LSD, DMT).

Table 4 summarizes the final hyperparameter configurations that yielded the best-performing models, along with their corresponding average custom scores. Table 5 presents the GPT-4 evaluation scores (1–5 scale) for each model across the five criteria, with averaged Overall Scores reported per substance. Furthermore, Table 5 reports GPT-4 rubric scores (1–5) per substance and model.

**Table 4:** Optimal hyperparameter settings with average Custom Score on test data.

| Model | Hyperparameters | Avg. Custom Score |
| --- | --- | --- |
| LexRank | compression_ratio = 0.30; similarity_threshold = 0.15; damping_factor = 0.85 | 0.46 |
| LSA | n_components = 24; min_cluster_size = 7; min_samples = 1; top_k_ratio = 0.25 | 0.39 |
| SBERT | relevance_weight = 0.6; diversity_weight = 0.2; position_bias = 0.05; similarity_threshold = 0.25; agg = “first” | 0.86 |

**Table 5:** GPT-4 evaluation scores across criteria (1–5) and Overall.

| Substance | Model | Coverage | Coherence | Clinical | Compression | Experience | Overall |
| --- | --- | --- | --- | --- | --- | --- | --- |
| DMT | LSA | 2.65 | 3.20 | 2.80 | 4.95 | 2.50 | 3.20 |
| DMT | LexRank | 3.05 | 3.20 | 3.35 | 4.65 | 3.35 | 3.70 |
| DMT | SBERT | 4.55 | 2.00 | 4.60 | 2.65 | 4.45 | 3.85 |
| LSD | LSA | 3.30 | 3.20 | 3.55 | 4.80 | 3.35 | 3.55 |
| LSD | LexRank | 3.20 | 3.10 | 3.25 | 4.80 | 3.45 | 3.45 |
| LSD | SBERT | 4.35 | 2.30 | 4.60 | 2.60 | 4.60 | 3.85 |
| Psilocybin | LSA | 3.15 | 3.20 | 3.30 | 4.75 | 3.10 | 3.50 |
| Psilocybin | LexRank | 3.25 | 3.00 | 3.25 | 4.85 | 3.30 | 3.50 |
| Psilocybin | SBERT | 4.25 | 2.40 | 4.40 | 2.75 | 4.55 | 3.85 |

Figure 2 presents radar chart visualizations, comparing the relative strengths and weaknesses of each model across evaluation dimensions by substance. SBERT showed high content coverage and experiential preservation but lower coherence, while LexRank maintained moderate scores across dimensions, and LSA excelled primarily in compression quality. Table 6 reports the TOPSIS closeness coefficients, synthesizing the multi-criteria evaluation scores into a ranked perspective across substances as well as overall. The results indicate that LexRank achieved the highest overall score, followed by SBERT and then LSA.

**Table 6:** TOPSIS closeness coefficients (0–1) across models and substances under equal weights.

| Substance | LexRank | LSA | SBERT |
| --- | --- | --- | --- |
| DMT | 0.53 | 0.43 | 0.57 |
| LSD | 0.48 | 0.53 | 0.51 |
| Psilocybin | 0.64 | 0.58 | 0.42 |
| Overall | 0.54 | 0.49 | 0.51 |

**Table 7:**
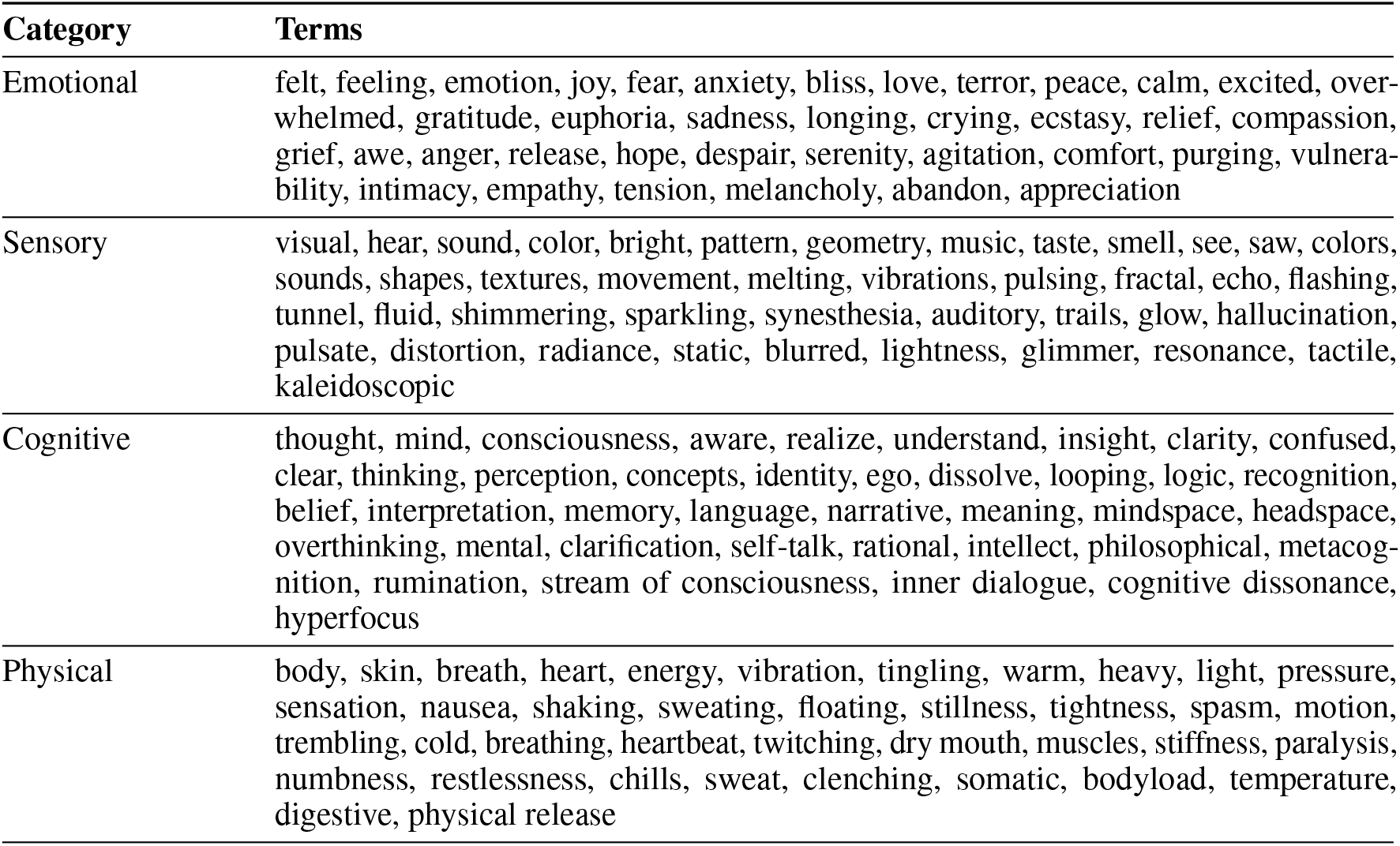
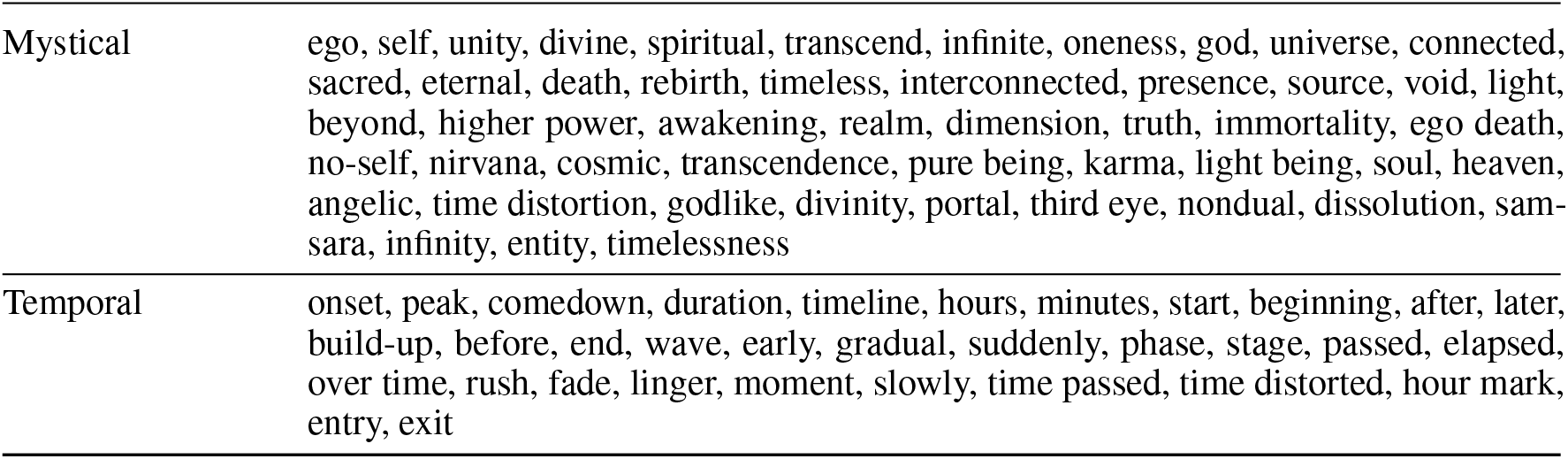
Full experiential lexicon categorized into emotional, sensory, cognitive, physical, mystical, and temporal domains.

**Figure 2.**
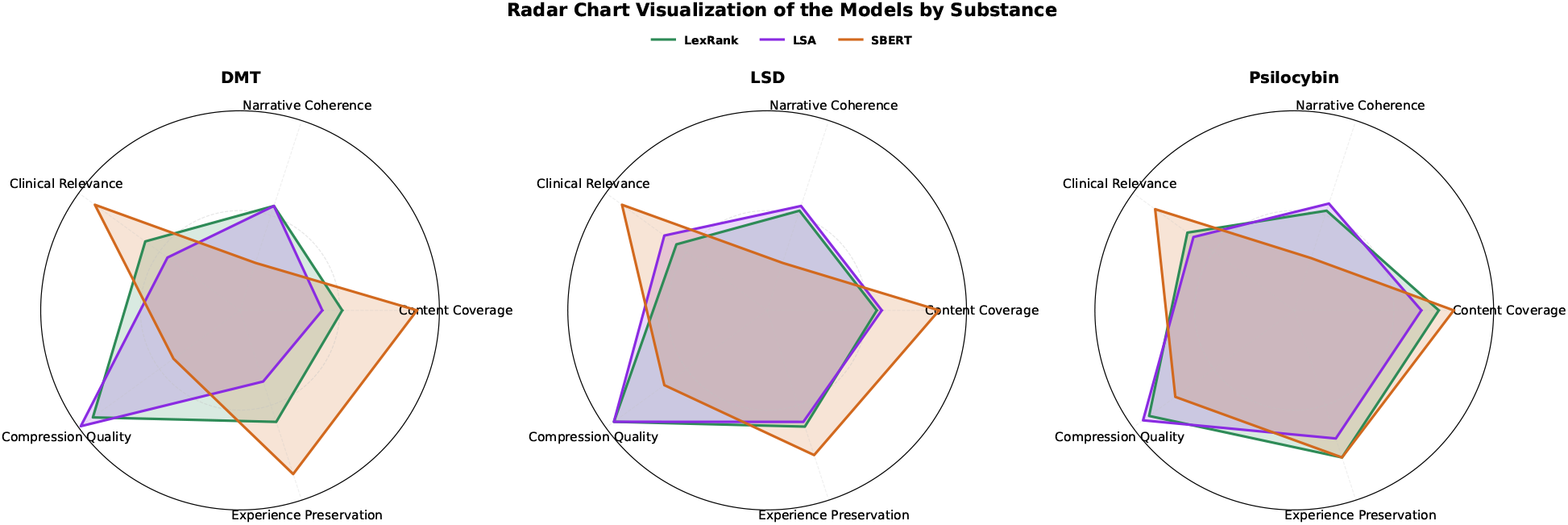
Radar chart visualization of model performance by substance across five criteria.

## 4 Discussion

Across substances, the three extractive pipelines showed complementary strengths that mirror well-documented trade-offs in clinical summarization. SBERT generally achieved higher LLM-based scores on content coverage, clinical relevance, and experiential preservation, though this often came at the expense of narrative coherence and length control, leading to longer or less organized summaries. LexRank and LSA, in contrast, tended to produce shorter and more fluent outputs with stronger coherence, but more frequently omitted dosing details, time course, or nuanced phenomenological content. This pattern echoes findings from medical text summarization research where content richness and readability often pull in opposite directions [26, 27].

When we aggregated the five evaluation dimensions with equal weights using TOPSIS, the results emphasized balance rather than maximal content capture. LexRank achieved the highest overall closeness score (0.54), followed by SBERT (0.51) and LSA (0.49). These differences highlight how modest deficits in coherence and compression can lower composite rankings even when content retention and clinical relevance are strong. Importantly, the rankings produced by the LLM rubric and TOPSIS should be seen as complementary: the former reveals performance on individual criteria, while the latter captures holistic balance across them [28].

Substance-specific patterns also suggest that no single method consistently dominates. SBERT performed better on dense and phenomenology-rich DMT narratives, where its embeddings captured subjective and temporal detail that rule-based methods tended to miss. LexRank was comparatively stronger on psilocybin reports, where recurring thematic motifs and structural coherence favored sentence-graph centrality. LSD reports, often shorter and less structurally complex, narrowed the gap between methods and allowed LSA to perform adequately despite its lower experiential retention. These variations indicate that model choice may depend as much on the structure and phenomenology of the source material as on overall algorithmic design.

From a practical standpoint, SBERT appears more suitable when completeness and clinical traceability are prioritized, such as in safety reviews or research coding. However, a lightweight post-processing stage that enforces target length, removes redundancies, and improves ordering could address its coherence limitations. LexRank is more appropriate for dashboards or registries where brevity and readability are essential, and it may also serve as a trimming layer for embedding-based methods. LSA remains a viable baseline when brevity and coherence are emphasized, although it risks losing important subjective detail in longer or more phenomenological narratives. These recommendations align with clinical NLP findings in radiology, discharge summary generation, and patient information summarization, where extraction and abstraction are often combined to balance factuality, readability, and coverage [29, 30].

Finally, the evaluation framework itself warrants reflection. Using GPT-4 as a calibrated rater under a fixed rubric proved useful for distinguishing between models. However, like other LLM-as-judge settings, it remains sensitive to prompt design and length effects [31]. Complementing this approach with TOPSIS helped make explicit the assumptions behind weighting choices and showed how rankings can shift depending on the balance of dimensions considered. Our study also has several limitations that should be noted. The evaluation was based on a relatively small sample of reports, which restricts generalizability. We focused exclusively on extractive pipelines and did not test abstractive summarization methods, which may offer different advantages and challenges. The absence of a standardized reference dataset limited our ability to benchmark against prior work. Our scoring function and weighting scheme, while transparent, represent only one possible configuration, and alternative formulations may yield different outcomes. Finally, relying on GPT-4 as a rater raises questions of reproducibility and potential bias.

Despite these constraints, the findings suggest clear directions for future work. Larger and more diverse collections of psychedelic reports would allow for more robust evaluation. Human adjudication could be incorporated alongside LLM judgments to strengthen reliability. Systematic testing of alternative scoring functions and weighting schemes would clarify how evaluation choices shape rankings. Abstractive and hybrid extraction–abstraction models deserve further exploration, as they may better capture the balance between factual coverage and narrative coherence. Taken together, these directions can help advance toward summarization systems that are both scientifically rigorous and clinically useful.

## Data Availability

We collected 400 narrative reports each for DMT, psilocybin, and LSD (total 1,200 reports) from the Erowid experience archive, with permission from the copyright holders.

https://www.erowid.org/

## Data Availability

The full-text reports are hosted by Erowid and cannot be redistributed due to copyright restrictions. Preprocessing code, analysis scripts, results and a representative example of generated summaries are available at: https://github.com/Shahidul2/Psychedelics-summary.

## Author Contributions

S.I. conceived the study, analyzed data, and interpreted results. S.S. collected data and conducted the literature review. M.N.H. supervised the study and revised the manuscript. All authors contributed equally to writing.

## Ethics Statement

This research complies with ethical publication standards and institutional review requirements. The study did not involve human subjects research, animal experimentation, or the collection of sensitive personal data. All source materials consisted of publicly available, fully anonymized narrative reports from Erowid, accessed with permission from the site administrators. A NER pipeline removed any residual personal identifiers to ensure total de-identification.

## A Full Experiential Lexicon

